# Performance of Elecsys pTau217 plasma immunoassay to detect brain amyloid pathology

**DOI:** 10.1101/2025.08.08.25333024

**Authors:** Derrek P. Hibar, Alina Bauer, Christina Rabe, Niels Borlinghaus, Alexander Jethwa, Gwendlyn Kollmorgen, Annunziata Di Domenico, Henrik Zetterberg, Kaj Blennow, Colin L. Masters, Reisa A. Sperling, Tobias Bittner

## Abstract

**INTRODUCTION:** We evaluated clinical performance of the fully-automated, high-throughput, prototype Elecsys^®^ Phospho-Tau (217P) plasma immunoassay (Roche Diagnostics) for detecting amyloid pathology.

**METHODS:** Plasma pTau217 levels were determined in samples from cognitively impaired and unimpaired individuals from five cohorts (*N*=7252) using the pTau217 plasma immunoassay. Clinical performance was evaluated against amyloid positron emission tomography.

**RESULTS:** For cognitively impaired, mean plasma pTau217 levels for amyloid-positive (A+) were higher (*n*=394; 0.835 pg/mL) than amyloid-negative (A–) individuals (*n*=144; 0.361 pg/mL); similarly, for cognitively unimpaired, A+ (*n*=224; 0.516 pg/mL) and A– individuals (*n*=1386; 0.220 pg/mL). Area under curve was 0.878 (95% confidence interval [CI] 0.840, 0.915) (impaired) and 0.907 (95% CI 0.885, 0.929) (unimpaired). Cutoff < 0.189 pg/mL reliably ruled-out individuals without amyloid pathology. High negative predictive values [92.51% (impaired); 98.60% (unimpaired)] were observed with sensitivity/specificity of 98.98%/29.17% and 95.54%/50.72%, respectively.

**DISCUSSION:** The pTau217 plasma immunoassay accurately detects amyloid pathology, irrespective of cognitive status.

**RESEARCH IN CONTEXT:** *1. Systematic review:* Prior literature on blood-based biomarkers, particularly pTau217, was reviewed using PubMed and recent conference proceedings. Several studies support plasma pTau217 as a promising biomarker to aid Alzheimer’s disease (AD) diagnosis, showing high concordance with amyloid positron emission tomography (PET) and cerebrospinal fluid biomarkers.

*2. Interpretation:* The high-throughput, fully automated Elecsys^®^ prototype pTau217 plasma immunoassay (Roche Diagnostics Ltd) accurately detected amyloid pathology across cognitively diverse populations in a large multi-cohort study (*N*=7252). The assay achieved a high area under curve (≥ 0.878) with respect to amyloid PET. A low-risk cutoff (< 0.189 pg/mL) showed good negative predictive value, supporting utility as a rule out tool.

*3. Future directions:* These results confirm the pTau217 plasma immunoassay as a reliable, scalable, and clinically useful biomarker for detection of amyloid pathology. Prospective validation in diverse populations and evaluation of outcomes in early AD care pathways are essential.

**Highlights:**

- Evaluated Elecsys^®^ pTau217 plasma immunoassay (Roche Diagnostics) in five cohorts.
- High performance detecting amyloid pathology in cognitively impaired/unimpaired.
- High negative predictive value supported utility as a pre-screener in AD.
- Identified cutoffs suitable as rule out pre-screener tools in clinical trials.
- Reinforced use of high-throughput pTau217 plasma measurements to aid AD diagnosis.

**Trial registration number:** A4, NCT02008357; SKYLINE, NCT05256134; AIBL, SAGE Project ID Number: 2022/PID06188; SVHM Local Ref ID: HREC 028/06; CREAD, NCT02670083; CREAD2, NCT03114657

## 1 BACKGROUND

Alzheimer’s disease (AD) is a progressive neurodegenerative disorder that is the leading cause of dementia and age-related cognitive and functional impairment.^1,2^ AD was ranked the seventh leading cause of death in the United States of America (USA) in the years 2020 and 2021.^1^ It is estimated that there are 50 million people living with AD in the USA.^2^ AD is characterized by the accumulation of amyloid-beta (Aβ) plaques and tau tangles in the brain, which lead to neuronal degeneration and cognitive decline.^1^

Recent biomarker evidence suggests that the underlying amyloid pathology associated with AD begins to be detectable around ≥ 20 years before the onset of clinical symptoms, which provides a window of opportunity for early diagnosis and therapy.^1^ Tools approved to aid AD diagnosis assess the underlying amyloid pathology, such as amyloid-positron emission tomography (PET) or the measurement of amyloid and tau proteins in cerebrospinal fluid (CSF).^3^ Amyloid PET visual read is considered the gold standard for determining amyloid positivity status^4^; however, many research studies use an exploratory quantitative method, the centiloid (CL) scale, which is considered a more standardized and accurate measure of amyloid burden across PET tracers.^5^ CSF-based diagnostic methods for amyloid pathology have shown high concordance with amyloid PET.^6^

There are a few disadvantages with the currently approved tools for detection of amyloid pathology; amyloid PET scans are expensive, expose patients to radiation and are not readily available in remote areas, and lumbar puncture as the method of choice to collect CSF may be perceived as invasive.^7^ With the recent approval of disease-modifying therapies (DMTs), such as donanemab and lecanemab,^8–11^ for the treatment of patients with early AD in several countries, there is a great need for a cost-effective, minimally invasive, and globally scalable method to identify those who could benefit from approved treatments as early in their disease course as possible. There is an anticipated surge in elderly individuals seeking AD diagnosis due to the expected widespread availability of these DMTs.^12^ According to the World Alzheimer Report, approximately 77% of multidisciplinary clinicians would be interested in new blood tests to improve diagnostic precision, especially with the expected increase in demand for AD diagnosis.^13^

Blood-based biomarkers (BBBMs) have been proposed as minimally invasive and cost-effective tools to rule in individuals who have a high likelihood of being amyloid-positive, and to rule out individuals who have a high likelihood of being amyloid-negative as measured by amyloid PET or CSF analysis.^14^ Plasma tau phosphorylated at threonine 217 (pTau217) is markedly increased in early- and late-stage AD, while levels are normal in most other neurodegenerative disorders.^15,16^ Thus, plasma pTau217 shows promise as a high-performing biomarker that can accurately rule in and rule out individuals with and without amyloid pathology, respectively.^15^ The prototype Elecsys^®^ Phospho-Tau (217P) Plasma immunoassay (Roche Diagnostics International Ltd, Rotkreuz, Switzerland) recently received US Food and Drug Administration (FDA) breakthrough device designation for the quantification of plasma pTau217, as an aid in identifying amyloid pathology.^17^

This study assessed the clinical performance of the prototype pTau217 plasma immunoassay for the detection of amyloid pathology with respect to amyloid PET visual read and different CL classifications across five clinical cohorts, comprising both cognitively impaired and unimpaired individuals with symptomatic or pre-symptomatic AD, as well as individuals without amyloid pathology.

## 2 METHODS

### 2.1 Retrospective study cohorts

Plasma pTau217 was measured in previously collected samples from five independent clinical cohorts comprising participants in the symptomatic or pre-symptomatic stages of AD, as well as individuals without amyloid pathology^18^: Anti-Amyloid Treatment in Asymptomatic Alzheimer’s Disease (A4) study (clinical data were downloaded from the Laboratory for Neuro Imaging Image and Data Archive [ida.loni.usc.edu] on April 15, 2020)^19^; SKYLINE^20^; Australian Imaging Biomarkers and Lifestyle (AIBL)^21^; CREAD^22^; and CREAD2.^22^

For A4, SKYLINE, CREAD, and CREAD2, samples from participants who were randomized into interventional studies based on a positive amyloid PET result were measured; samples from participants who failed screening based on a negative amyloid PET result and therefore not randomized into interventional studies, were also measured. This allowed for the comparison of plasma pTau217 in amyloid PET-positive and -negative individuals within the context of these studies. Additionally, SKYLINE was terminated prematurely, resulting in a limited sample size; consequently, results are only available for participants who progressed far enough in the screening process prior to discontinuation.

A4 and SKYLINE comprised cognitively unimpaired individuals, while AIBL consisted of both cognitively impaired and unimpaired individuals, and CREAD and CREAD2 only included cognitively impaired individuals. Participants across the five cohorts (including screen failures) were selected based on the following criteria: aged ≥ 50 and ≤ 90 years; and the availability of (i) amyloid PET results, (ii) plasma samples collected within 12 months before or after the most recent amyloid PET scan, and (iii) plasma pTau217 data.

A4 was a multi-center study across Australia, Canada, Japan, and the USA; AIBL was conducted at two sites in Australia; and SKYLINE, CREAD, and CREAD2 were global, multi-center studies.^19–22^ Participants enrolled in the A4 study were aged 65–85 years, living independently, and had a Clinical Dementia Rating-Global Score (CDR-GS) of 0 (cognitively unimpaired), a mini-mental state examination score of ≥ 25, and elevated brain amyloid levels detected by quantitative 18F-florbetapir PET imaging.^19^ For A4, amyloid positivity was primarily determined using quantitative measures of standardized uptake value ratio (SUVr) ≥ 1.15, with visual read results only utilized at specific quantitative measurement levels. This approach means that a subset of individuals was classified as amyloid-positive based on quantitative thresholds, despite having negative visual reads.^19^ However, the main analysis of A4 presented herein utilized the amyloid PET visual read findings. The SKYLINE study comprised participants aged 60–80 years, who were cognitively unimpaired, determined to have CDR-GS of 0 and Repeatable Battery for the Assessment of Neuropsychological Status Delayed Memory Index of ≥ 80. Amyloid-positive individuals were enrolled determined by either a positive amyloid PET visual read or CSF tau phosphorylated at threonine181/Aβ 1–42 (pTau181/Aβ42) ratio of > 0.04.^20^ The AIBL study enrolled participants aged ≥ 60 years who had AD dementia, mild cognitive impairment, or were cognitively unimpaired.^21^ Both CREAD and CREAD2 enrolled participants aged 50–85 years with prodromal or mild AD consistent with National Institute on Aging–Alzheimer Association criteria, amyloid positivity determined by CSF-based results or available amyloid PET visual read, a mini-mental state examination score of ≥ 22, a CDR-GS of 0.5 or 1.0, and abnormal memory function defined by a Free and Cued Selective Reminding Test cueing index score of ≤ 0.67 and a free recall score of ≤ 27.^22^

### 2.2 Plasma pTau217 measurement

Plasma pTau217 was measured using the prototype pTau217 plasma immunoassay on the high-throughput Cobas^®^ e 801 analytical unit as part of the NeuroToolKit study setup (both Roche Diagnostics International Ltd, Rotkreuz, Switzerland). The prototype immunoassay used in this study is a newer version (developed in 2024)^23,24^ compared with that used in previously published studies from 2023,^25,26^ with considerably improved analytical sensitivity, i.e., lower limit of quantification. Biomarker concentrations were measured from samples immediately after thawing in several batches at one laboratory per study (A4 at Labcorp TBS [Indianapolis, Indiana, USA]; SKYLINE at Labcorp Central Laboratory Services [Indianapolis, Indiana, USA]; CREAD and CREAD2 at University of Gothenburg [Mölndal, Sweden]; AIBL at Microcoat [Bernried, Germany]). Absolute biomarker concentrations (i.e., pg/mL values) are subject to change due to the prototype status of the assay.

Samples from A4 were collected as previously described^27^ and plasma pTau217 was measured in leftover samples that were previously thawed and frozen at least one more time. Blood samples from SKYLINE were collected into 2×10 mL K2-ethylenediaminetetraacetic acid tubes (BD366643, Becton Dickinson, Franklin Lakes, New Jersey, USA). Tubes were inverted 8–10 times and within 60 minutes centrifuged at 1500×g at 4°C for 10 minutes. A total of 0.5 mL of blood plasma was aliquoted into 0.5 mL tubes (72.730.005, Sarstedt, Nümbrecht, Germany) and immediately frozen at −70°C. Blood samples from AIBL were collected as previously described^21^ and pTau217 was measured in leftover samples that were previously thawed and frozen at least one more time. Blood samples from CREAD and CREAD2 were collected as previously described.^22^ Based on prior data, up to two freeze/thaw cycles are acceptable for Elecsys pTau217 and do not cause any significant changes in pTau217 levels in a sample.^26^

### 2.3 Amyloid PET

In this retrospective study, amyloid PET scans were retrieved from various databases linked to the original studies. Amyloid positivity was determined by amyloid PET visual read as per the package inserts of the three FDA-approved PET tracers: Amyvid,^28^ Vizamyl,^29^ and Neuraceq^30^ as previously described for each study in section 2.1. Prototype pTau217 plasma immunoassay results were also compared with CL classifications of amyloid positivity at selected cutoffs of ≥ 24 and ≥ 40. For this analysis, A4 florbetapir SUVr values were converted to CL following the procedure in Navitsky et al. 2018.^31^ AIBL multi-tracer SUVr data were converted to CL following the procedures outlined in Bourgeat et al. 2018.^32^ CL values from multi-tracer SUVr data in SKYLINE, CREAD, and CREAD2 were calculated using image analysis pipelines calibrated according to the procedure detailed by Klunk et al. 2015.^33^

### 2.4 Statistical analysis

Baseline characteristics were demonstrated using descriptive analyses. Receiver operating characteristic (ROC) area under curve (AUC) analyses were performed to evaluate the overall discriminative ability of plasma pTau217. Forest plots were generated to show the AUC results within cognitively impaired and unimpaired cohorts and across individual studies. Integrated risk profiling was used to determine the performance of plasma pTau217 over the full range of possible cutoffs for identifying cognitively impaired and unimpaired individuals with underlying amyloid pathology (including amyloid-positive and amyloid-negative individuals). Integrated risk profiling included: i) integrated risk plots that provide a unified visual representation of the performance in practice based on predictiveness curves,^34^ incorporating estimated prevalences (including the ability to specify adjusted prevalences) to view key performance metrics including the negative predictive value (NPV; sometimes reported as 1-NPV), positive predictive value (PPV), and screen-out rates for all possible cutoffs; and ii) integrated sensitivity and specificity plots. Performance metrics were calculated for amyloid status classification with amyloid PET visual read, CL ≥ 24, and CL ≥ 40.

We evaluated the suitability of the plasma pTau217 immunoassay for use as a pre-screening tool to increase the efficiency of enrollment in a theoretical clinical trial of early symptomatic or pre-symptomatic AD seeking to enroll 750 individuals. This scenario assumed that individuals referred to the clinical trial would be a mixture of cognitively impaired and cognitively unimpaired participants, but the exact mixture was unknown, and the true cognitive status was not available at the time of pTau217 assessment. We used integrated risk profiling to identify a cutoff of plasma pTau217 concentration and provide estimates of performance ranges, efficiency, and cost savings when used for pre-screening trial enrollment.^35^ All analyses were conducted in R (v4.3.1) and the integrated risk profiling was performed using tools developed as part of the stats4phc R package (v0.1.2).^36^

## 3 RESULTS

### 3.1 Baseline characteristics

In total, 7252 individuals were eligible for inclusion in the study across the five clinical cohorts; 1720 individuals were cognitively impaired and 5532 were cognitively unimpaired participants. Of these participants, 538 cognitively impaired and 1610 cognitively unimpaired individuals had available plasma pTau217 measurements; the remaining participants were excluded for various reasons (sample volume constraints, availability of amyloid PET, or pTau217 measurements). The subset of participants with available plasma pTau217 measurements (Table 1) demonstrated baseline characteristics generally comparable with those of the overall enrolled population (Table S1 in the supporting information).

**TABLE 1.**
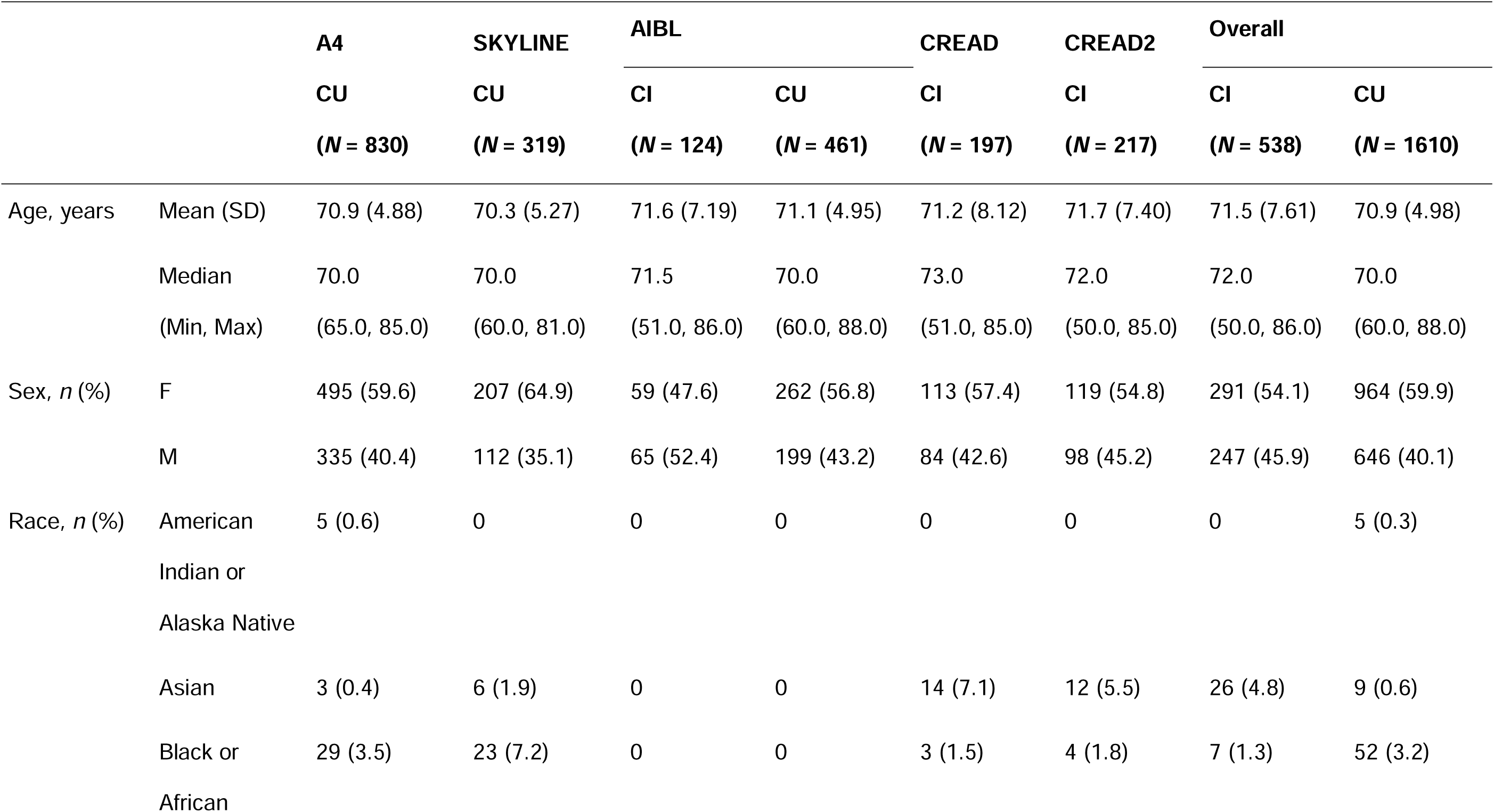
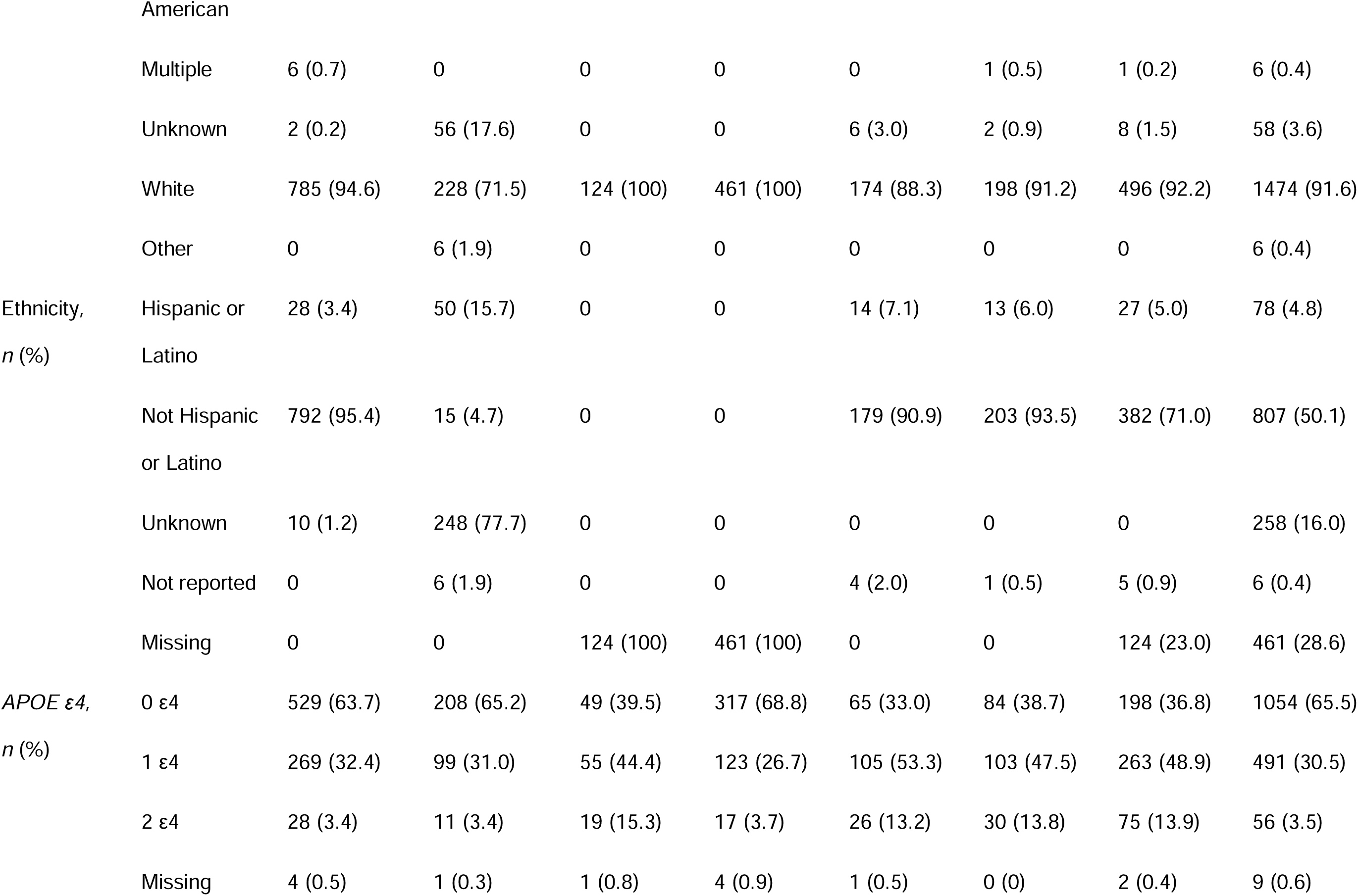
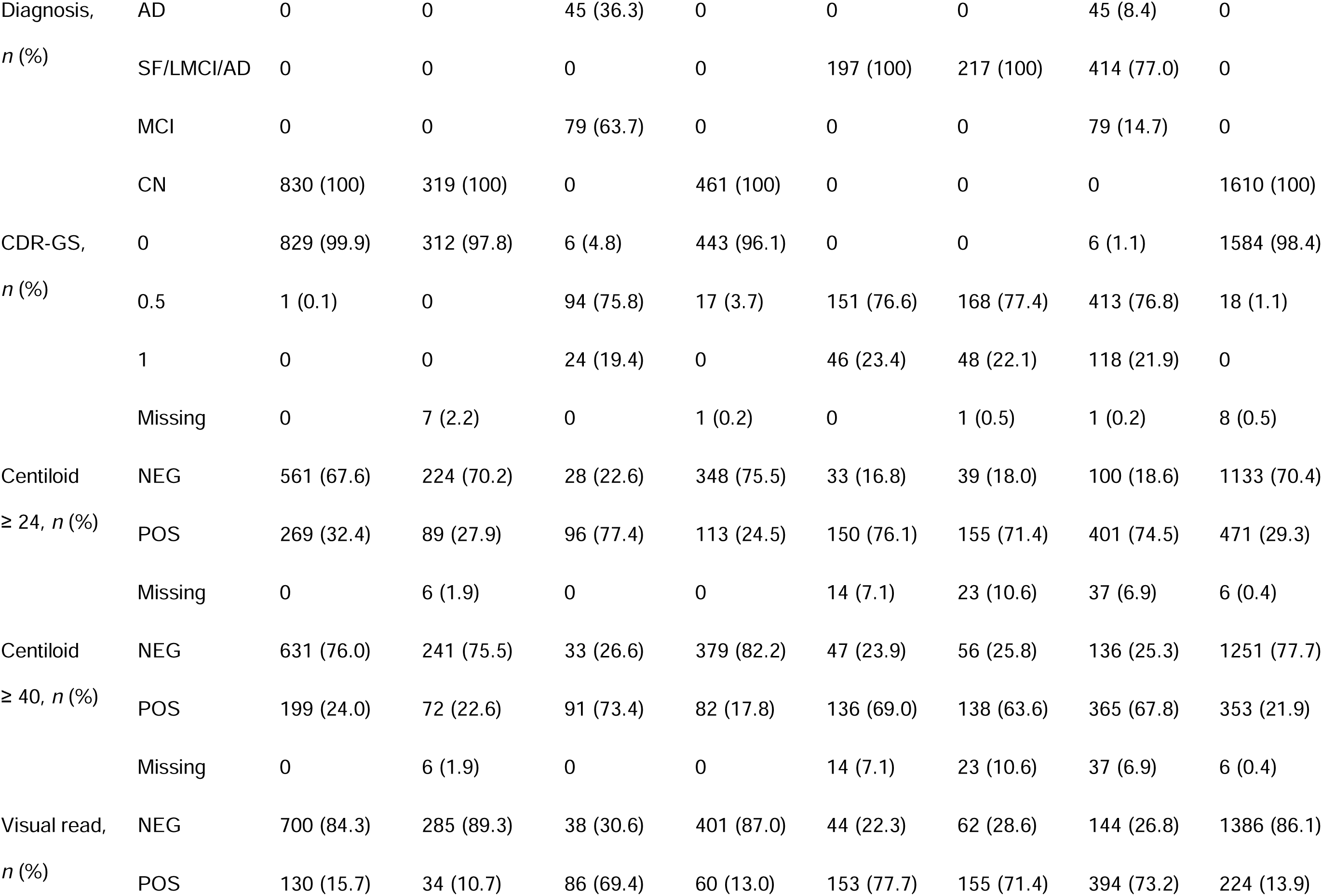
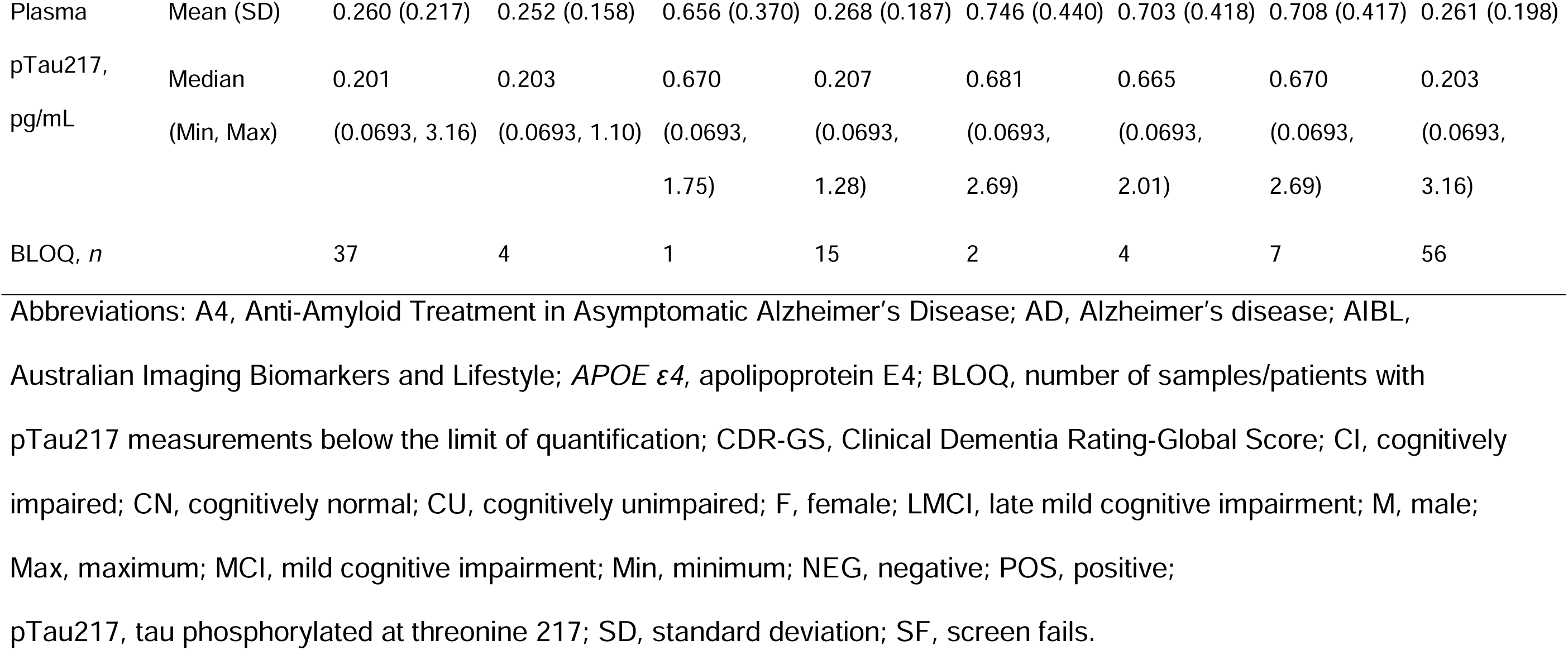
Summary of baseline characteristics of eligible participants with an available plasma pTau217 measurement and amyloid PET visual read.

For the subset of participants with available plasma pTau217 measurements, the mean age (standard deviation [SD]) for cognitively impaired and unimpaired individuals was 71.5 (SD=7.61) years and 70.9 (SD=4.98) years, respectively. The proportions of female cognitively impaired versus cognitively unimpaired individuals were 54.1% and 59.9%, respectively. The majority of both cognitively impaired (92.2%) and unimpaired (91.6%) individuals were White. Baseline characteristics across cohorts were largely comparable within the groups of cognitively impaired and unimpaired individuals; except for differences in race and ethnicity, which can be attributed to variation in geographic locations across the cohorts.

Across all participants with available pTau217 measurements, the observed differences between cognitively impaired and unimpaired individuals were consistent with our expectations, for instance the higher prevalence of apolipoprotein E4 (*APOE* ε*4*) carriers in the cognitively impaired compared with unimpaired individuals (Table 1). Further, and as expected, 98.7% of cognitively impaired individuals had a CDR-GS of ≥ 0.5 compared with 1.1% of cognitively unimpaired/normal individuals, while the amyloid positivity prevalence using amyloid PET visual read was 73.2% and 13.9% in cognitively impaired and unimpaired individuals, respectively, across all individuals with available pTau217 measurements.

Amyloid positivity prevalence in the full set of cognitively impaired individuals was 69.9% (Table S1 in the supporting information). The availability of plasma samples for testing was somewhat limited in the screen failures in the CREAD studies. The amyloid positivity prevalence in the full study was more representative and, therefore, the adjusted reported statistics for the cognitively impaired population used the 69.9% prevalence instead of the 73.2% prevalence. Similarly, the prevalence of amyloid PET positivity at selected CL cutoffs of ≥ 24 and ≥ 40 was higher in cognitively impaired individuals (≥ 24 CL cutoff: 77.9%; ≥ 40 CL cutoff: 70.5%) compared with cognitively unimpaired individuals (≥ 24 CL cutoff: 29.3%; ≥ 40 CL cutoff: 22.0%). Statistics reported for the ≥ 24 CL and ≥ 40 CL cutoffs in cognitively impaired individuals used the amyloid positivity prevalence in the full cohort and were 77.3% and 70.3%, respectively.

### 3.2 Performance of the prototype pTau217 plasma immunoassay across the five clinical cohorts

#### 3.2.1 Distribution of plasma pTau217 in relation to amyloid PET visual read

Within cognitively impaired individuals, the mean plasma pTau217 level of amyloid-positive individuals was higher (0.835 pg/mL [SD=0.381]) relative to amyloid-negative individuals (0.361 pg/mL [SD=0.294]) (Table S2 in the supporting information), a trend observed across individual cohorts (Figure 1). We observed a similar elevation in mean pTau217 in cognitively unimpaired amyloid-positive individuals (0.516 pg/mL [SD=0.235]) relative to amyloid-negative individuals (0.220 pg/mL [SD=0.157]) (Table S2 in the supporting information), which was consistent across individual cohorts (Figure 1). In addition, mean (SD) plasma pTau217 levels of cognitively impaired individuals (0.708 pg/mL [SD=0.417]) were higher compared with cognitively unimpaired individuals (0.261 pg/ mL [SD=0.198]) regardless of amyloid status (Table 1). Out of 2148 measured samples, 63 (2.93%) were below the limit of quantification.

**FIGURE 1.**
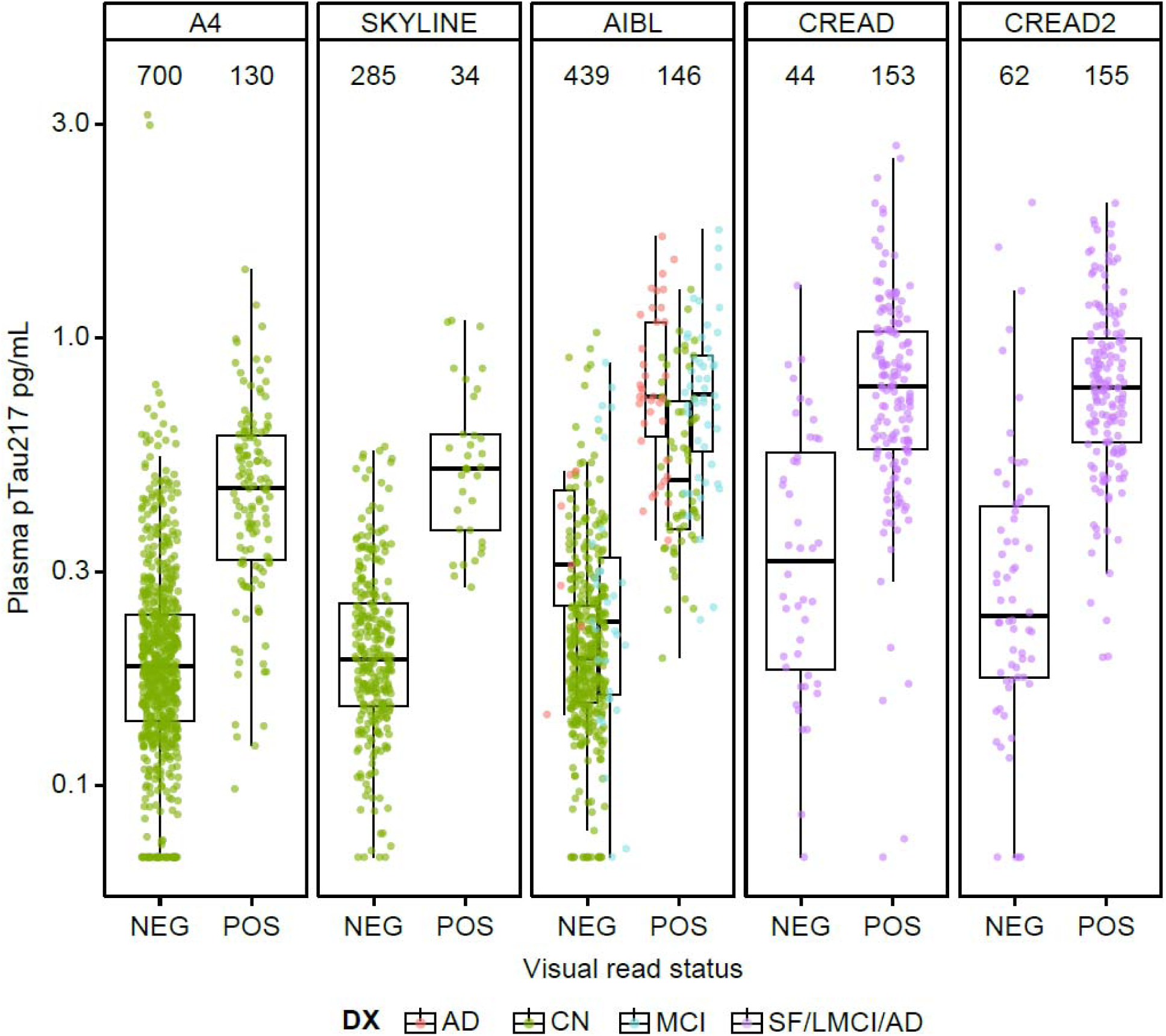
Plasma pTau217 levels by amyloid PET visual read status further split by diagnosis across the five clinical cohorts. Abbreviations: A4, Anti-Amyloid Treatment in Asymptomatic Alzheimer’s Disease; AD, Alzheimer’s disease; AIBL, Australian Imaging Biomarkers and Lifestyle; CN, cognitively normal; DX, diagnosis; LMCI, late mild cognitive impairment; MCI, mild cognitive impairment; NEG, negative; PET, positron emission tomography; POS, positive; pTau217, tau phosphorylated at threonine 217; SF, screen fails.

#### 3.2.2 Forest plots and ROC-AUC analyses with respect to amyloid PET visual read status

In cognitively impaired individuals, the combined AUC for plasma pTau217 with respect to amyloid PET visual read was 0.878 (95% confidence interval [95% CI] 0.840, 0915) across the cohorts. AUCs for individual cohorts were: 0.912 (95% CI 0.854, 0.970) (AIBL), 0.837 (95% CI 0.768, 0.907) (CREAD), and 0.889 (95% CI 0.828, 0.951) (CREAD2), as shown in Figure 2A and Figure S1A. In cognitively unimpaired individuals, a combined AUC of 0.907 (95% CI 0.885, 0.929) was observed across the cohorts for plasma pTau217 with respect to amyloid PET visual read. AUCs for individual cohorts were: 0.883 (95% CI 0.849, 0.917) (A4), 0.954 (95% CI 0.930, 0.978) (SKYLINE), and 0.935 (95% CI 0.909, 0.961) (AIBL), as shown in Figure 2B and Figure S1B.

**FIGURE 2.**
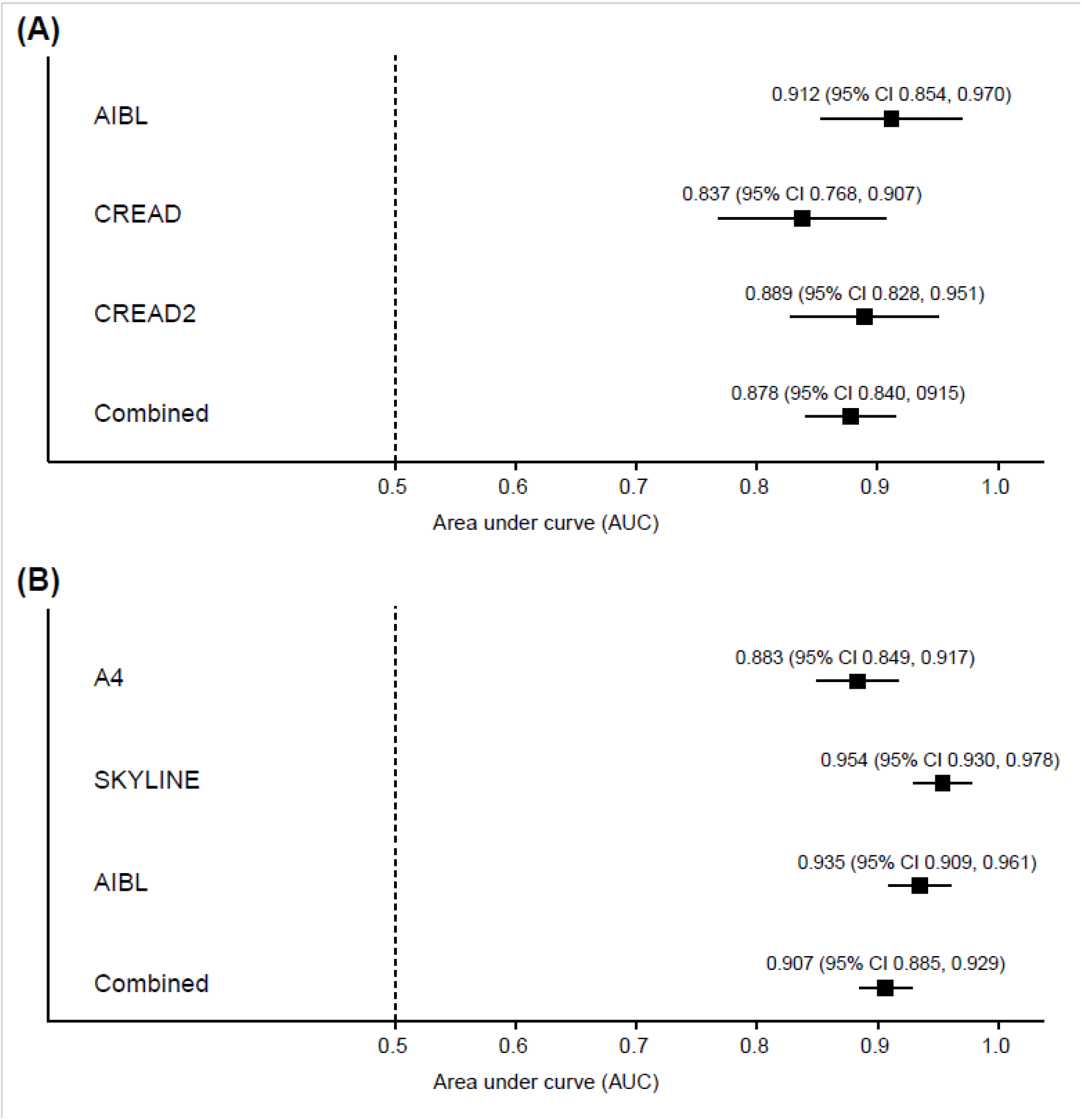
Forest plot analyses for plasma pTau217 with respect to amyloid PET visual read status in (A) cognitively impaired and (B) cognitively unimpaired individuals. Each black square represents an AUC estimate with the surrounding lines representing the 95% confidence interval. Abbreviations: 95% CI, 95% confidence interval; A4, Anti-Amyloid Treatment in Asymptomatic Alzheimer’s Disease; AIBL, Australian Imaging Biomarkers and Lifestyle; AUC, area under curve; PET, positron emission tomography; pTau217, tau phosphorylated at threonine 217.

#### 3.2.3 Performance as a pre-screening tool in trials enrolling based on amyloid PET visual read

Integrated risk plots for cognitively impaired (*n* = 538; Figure 3A) and cognitively unimpaired (*n* = 1610; Figure 3B) individuals show the PPV and 1-NPV over a full range of cutoffs of the prototype pTau217 plasma immunoassay. We chose a cutoff (< 0.189 pg/mL) with a low rate of amyloid PET visual read-positive individuals among those screened out of the study (i.e., a high NPV) in both the cognitively impaired and unimpaired population. The NPV for the prototype pTau217 plasma immunoassay at the selected < 0.189 pg/mL cutoff was 92.51% in the cognitively impaired population and 98.60% in the cognitively unimpaired population, respectively (Table 2). The corresponding sensitivity and specificity of the assay using a cutoff of < 0.189 pg/mL were 98.98% and 29.17% in the cognitively impaired population, and 95.54% and 50.72% in the cognitively unimpaired population, respectively (Figure 4). Further, we estimate that implementing the < 0.189 pg/mL cutoff as a pre-screening tool could reduce the total number of amyloid PET scans required for full study enrollment by 8.55% (cognitively impaired) and 41.68% (cognitively unimpaired), while requiring only a 1.03% (cognitively impaired) and 4.67% (cognitively unimpaired) increase in the total number of participants needing pre-screening (Table 2). We report performance characteristics covering the full range of potential cutoffs in cognitively impaired and unimpaired individuals to enable performance estimates for different cutoffs and use cases (Tables S3 and S4 in the supporting information).

**FIGURE 3.**
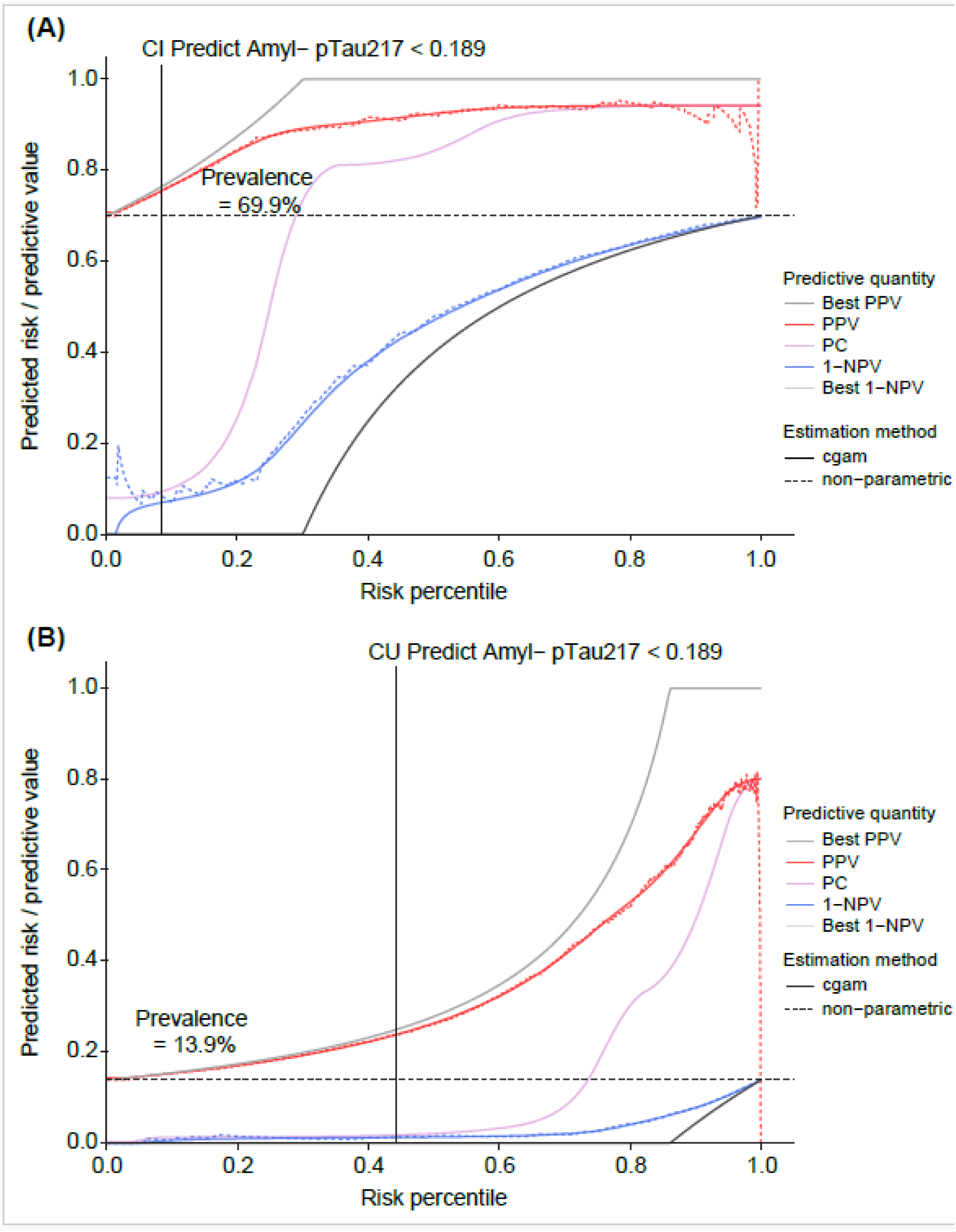
Integrated risk plots for (A) cognitively impaired and (B) cognitively unimpaired individuals. The red lines represent the PPV at a given risk percentile of pTau217 (for the smoothed cgam and non-parametric estimates). Similarly, the blue lines indicate the 1-NPV. The pink line that intersects the dotted midway line indicates predictiveness. The solid gray lines indicate the performance of a perfect assay applied to the same population as a reference. The dashed horizontal line indicates the prevalence of amyloid PET visual read positivity in that population. The black vertical line indicates the performance characteristics associated with the example rule out cutoff (0.189 pg/mL) at a given percentile of pTau217. Abbreviations: 1-NPV, 1 - negative predictive value; cgam, constrained generalized additive model; CI, cognitively impaired; CU, cognitively unimpaired; PC, predictiveness curve; PET, positron emission tomography; PPV, positive predictive value; pTau217, tau phosphorylated at threonine 217.

**FIGURE 4.**
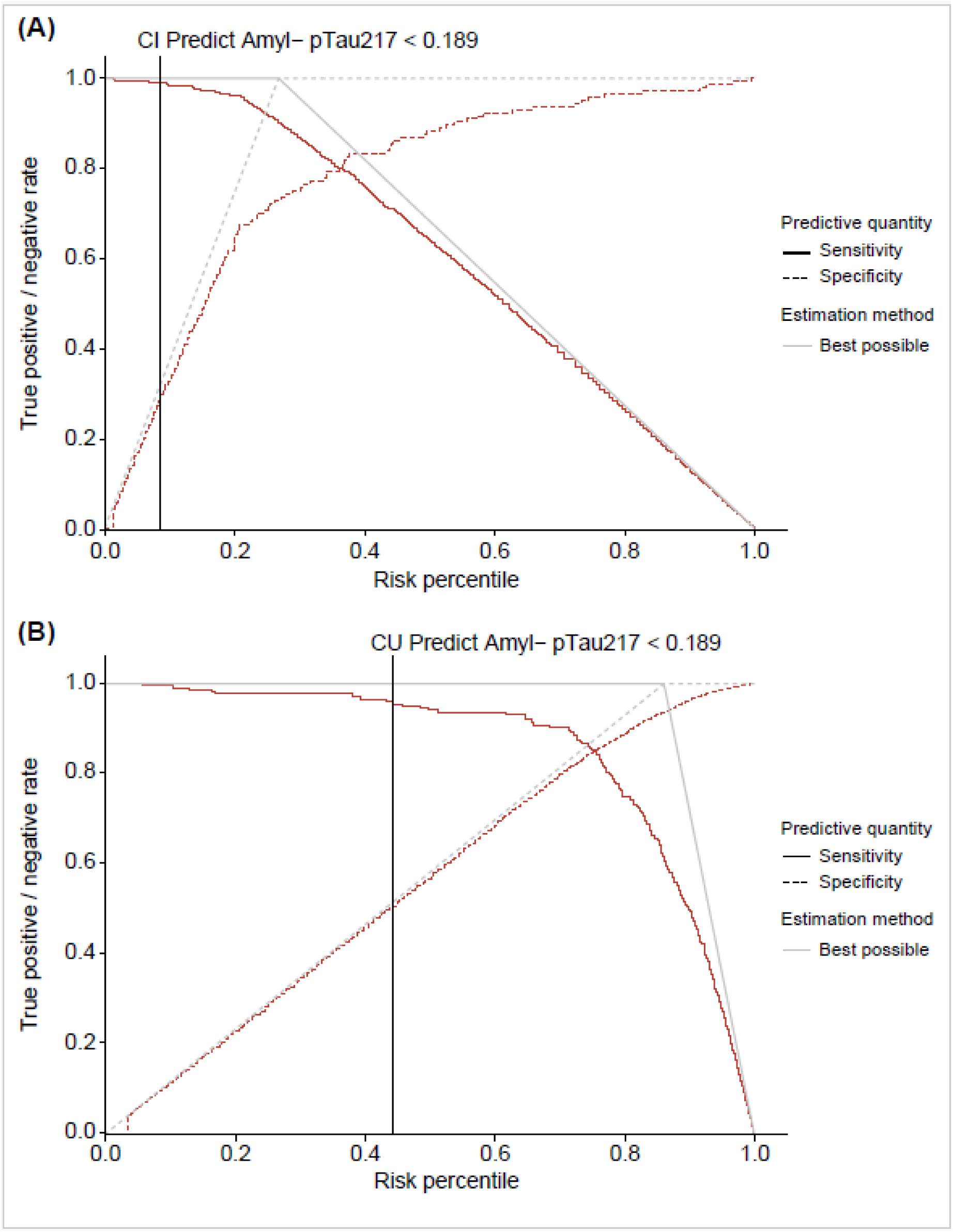
Sensitivity and specificity plots for (A) cognitively impaired and (B) cognitively unimpaired individuals. The red lines represent the sensitivity (solid line) and the specificity (dashed line) at a given risk percentile of pTau217. The solid gray lines indicate the performance of a perfect assay applied to the same population as a reference. The black vertical line indicates the performance characteristics associated with the example rule out cutoff (0.189 pg/mL) at a given percentile of pTau217. Abbreviations: CI, cognitively impaired; CU, cognitively unimpaired; pTau217, tau phosphorylated at threonine 217.

**TABLE 2.** Performance characteristics of the prototype pTau217 plasma immunoassay, using a cutoff of < 0.189 pg/mL, for pre-screening rule out of amyloid pathology determined by amyloid PET visual read in cognitively impaired and cognitively unimpaired individuals.

| **Performance characteristics** | **CI (n = 538)** | **CU (n = 1610)** |
| --- | --- | --- |
| Positive amyloid PET visual read, *n* (%) | 394 (73.2) | 224 (13.9) |
| Negative amyloid PET visual read, *n* (%) | 144 (26.8) | 1386 (86.1) |
| Prevalence, % | 69.94* | 13.91 |
| True-positive individuals, *n* (%) | 390 (72.49) | 214 (13.29) |
| False-negative individuals, *n* (%) | 4 (0.74) | 10 (0.62) |
| False-positive individuals, *n* (%) | 102 (18.96) | 683 (42.42) |
| True-negative individuals, *n* (%) | 42 (7.81) | 703 (43.66) |
| OPA, % | 80.30 | 56.96 |
| PPV, % | 76.48 | 23.86 |
| NPV, % | 92.51 | 98.60 |
| Sensitivity, % | 98.98 | 95.54 |
| Specificity, % | 29.17 | 50.72 |
| Screen-out rate, % | 9.48 | 44.29 |
| Additional screening required, % | 1.03 | 4.67 |
| Amyloid PET scans saved, % | 8.55 | 41.68 |

\*Note that the prevalence of amyloid positivity in the CI group is using the adjusted value of 69.9% measured in the full CI cohort.
Abbreviations: CI, cognitively impaired; CU, cognitively unimpaired; NPV, negative predictive value; OPA, overall percent agreement; PET, positron emission tomography; PPV, positive predictive value; pTau217, tau phosphorylated at threonine 217.

#### 3.2.4 Performance with respect to CL classifications

The optimal cutoff for the prototype pTau217 plasma immunoassay depends on the use case and population (e.g., cognitively impaired vs unimpaired individuals). Demographics for the subset of participants who had available CL and pTau217 plasma immunoassay measurements are summarized in Table S5 in the supporting information. The lower CL cutoff of ≥ 24 leveraged the quantitative nature of the CL metric to identify additional amyloid-positive individuals with lower amyloid burden and thus the prevalence of amyloid-positive individuals increased across all five clinical cohorts compared with amyloid PET visual read results (Figure S2). Despite the differences in defining amyloid positivity status, overall performance of the prototype pTau217 plasma immunoassay was consistent. When defining amyloid positivity with the ≥ 24 CL definition, we observed an AUC of 0.872 (95% CI 0.828, 0.917) in cognitively impaired and 0.864 (95% CI 0.842, 0.887) in cognitively unimpaired individuals. With the ≥ 40 CL definition, we observed an AUC of 0.849 (95% CI 0.807, 0.891) in cognitively impaired and 0.911 (95% CI 0.892, 0.930) in cognitively unimpaired individuals. We provide a full range of select cutoffs and associated performance metrics for the CL-based amyloid definitions to explore the performance of different cutoffs and enable different use cases for trial and study planning in Tables S6−9 in the supporting information.

## 4 DISCUSSION

This study assessed the clinical performance of the prototype pTau217 plasma immunoassay for ruling out amyloid pathology, in relation to amyloid PET visual read, across five clinical cohorts comprising cognitively impaired and unimpaired individuals with symptomatic or pre-symptomatic AD, as well as individuals without amyloid pathology. Enrolled individuals with available pTau217 measurements (*n* = 2148) demonstrated comparable baseline characteristics, differences reported between cognitively impaired and unimpaired individuals in terms of *APOE* ε*4* status, clinical diagnosis, and prevalence of amyloid positivity were in line with previous literature.^37^ Mean plasma pTau217 levels were elevated in amyloid-positive individuals relative to amyloid-negative individuals regardless of cognitive status, supporting the connection between plasma pTau217 and amyloid pathology as well as its use in both cognitively impaired and unimpaired individuals. The findings in this study demonstrate that the pTau217 plasma immunoassay is a robust and scalable diagnostic tool for excluding individuals who are unlikely to have amyloid pathology, with a particularly high NPV of 92.5% in cognitively impaired and 98.6% in cognitively unimpaired individuals at a potential pre-screening cutoff of < 0.189 pg/mL, supporting its utility as a reliable rule out tool. Taken together, these results promote the utility of the prototype pTau217 plasma immunoassay as a triaging diagnostic tool, reducing the reliance on invasive and costly approaches while supporting clinical implementation in AD research and clinical care pathways.

Guided by integrated risk profiling, we evaluated the performance of the plasma pTau217 assay for use as a pre-screening tool for a theoretical trial enrolling early AD participants. We sought to identify a cutoff that would exclude individuals who are unlikely to be amyloid-positive (i.e., not invited for further screening) in both cognitively impaired and unimpaired populations. Ideally, the cutoff would achieve a high NPV > 90% (or equivalently a 1-NPV of < 10%) while also maintaining a high screen-out rate (as close to the amyloid-negative prevalence as possible). In practice, this approach would enrich the screened-in population for amyloid positivity and directly reduce the total number of amyloid PET scans performed to achieve full study enrollment. We found that the prototype pTau217 plasma immunoassay demonstrated robust screen-out performance across the five clinical cohorts, with a PPV of 76.48%, NPV of 92.51%, sensitivity of 98.98%, and specificity of 29.17% in cognitively impaired individuals and a PPV of 23.86%, NPV of 98.60%, sensitivity of 95.54%, and specificity of 50.72% in cognitively unimpaired individuals, at a cutoff of < 0.189 pg/mL. The high sensitivities and high NPV regardless of cognitive status support the ability of the assay to accurately rule out amyloid-negative individuals. We also found that use of this assay could lead to significant savings in terms of patient recruitment times and reduce the total number of confirmatory amyloid PET scans required for trial enrollment. We estimate that implementing the plasma pTau217 immunoassay as a pre-screening tool at the cutoff of < 0.189 pg/mL could result in an 8.55% (cognitively impaired) and 41.68% (cognitively unimpaired) reduction in the total number of amyloid PET scans required for full enrollment. This is a significant saving in terms of time and cost to study teams, and reduces the burden on participants who ultimately are not eligible for the trial. The range of 8.55% to 41.68% is rather broad; however, the observed savings are heavily based on the patient population that enters the screening funnel and represents two distinct clinical scenarios. Lower savings (8.55%; cognitively impaired) are observed in symptomatic populations, where clinical pre-selection increases amyloid pathology prevalence; conversely, higher savings (41.68%; cognitively unimpaired) are seen in asymptomatic or unselected populations, where the biomarker’s high sensitivity effectively screens out more amyloid-negative individuals, thus reducing the need for unnecessary confirmatory PET scans. If the recruited patient population heavily comprises participants without cognitive impairment, the savings from including the assay as a pre-screening tool increase.^38^ We suspect this scenario can happen when recruiting for an early AD trial when patient recruitment happens in a community setting outside of specialized memory centers, where participants are more likely to have confirmed AD-related cognitive impairment diagnosis.

In addition, the prototype pTau217 plasma immunoassay showed high concordance with amyloid PET results visual read in cognitively impaired individuals (AUC=0.878) and cognitively unimpaired individuals (AUC=0.907) across cohorts. AUCs for individual cohorts were comparable with the overall ROC results. These findings align with results obtained in cognitively impaired individuals with another pTau217 immunoassay, the Lumipulse G pTau217 plasma immunoassay to identify amyloid pathology (AUC=0.862) by amyloid PET.^24^ The prototype pTau217 plasma immunoassay also demonstrated high concordance with CL classifications in cognitively impaired and unimpaired individuals, particularly at a cutoff of ≥ 40 (impaired: 0.849; unimpaired: 0.911) compared with a cutoff of ≥ 24 (impaired: 0.872; unimpaired: 0.864).

Although the five clinical cohorts originated from various geographic locations and included hundreds of clinical sites, and despite differences in preanalytical sample handling (including different freeze/thaw cycles), sample age, and numerous batches of measurements using multiple instruments across varied laboratories (referred to in section 2.2), the prototype pTau217 plasma immunoassay demonstrated robust performance regardless of cognitive status. This supports the potential clinical utility of the fully automated and high-throughput assay as a diagnostic tool for identifying amyloid-positive and amyloid-negative individuals with a high and low likelihood of developing AD, respectively, as an equivalent to amyloid PET visual read, the current gold standard.^4,17^ Plasma pTau217 as a BBBM has various advantages, namely, it is scalable, accessible, and more affordable compared with amyloid PET. In contrast, amyloid PET faces limitations such as restricted accessibility due to the low number of scanners and specialized personnel, as well as its higher cost. Moreover, although CSF analysis has proven valuable in AD diagnosis, it has limitations due to its perceived invasiveness. BBBMs address these challenges by enabling the detection of amyloid pathology at the earliest stages, providing an opportunity to intervene when patients are most likely to benefit from DMTs.^39^ Furthermore, BBBMs are minimally invasive alternatives to CSF analysis as well as cost-effective alternatives to amyloid PET imaging.^40^ With the anticipated surge of elderly individuals seeking AD diagnosis due to the availability of DMTs, BBBMs, in particular plasma pTau217, will be critical in enabling timely AD diagnosis and treatment.^12,13^ While mass spectrometry-based approaches are accurate,^41^ fully automated immunoassays offer several practical advantages over mass spectrometry, such as broader availability, reduced costs, and faster turnaround times,^42^ which further support the widespread utility in the clinical setting.

This study included a large sample size comprising participants from five clinical cohorts (*N* = 7252) in order to estimate the performance of the prototype pTau217 plasma immunoassay with high confidence. A limitation of this study was the lack of diversity in the study population with very few participants from underrepresented groups. Another limitation is the use of participants from memory center settings rather than from primary care settings. Further research is required to validate these findings prospectively in more diverse populations and define the potential role of the prototype pTau217 plasma immunoassay in routine clinical practice, including the primary care setting.

## CONCLUSIONS

The findings of this study demonstrate the robust performance of the prototype pTau217 plasma immunoassay in measuring plasma pTau217 as a biomarker, effectively detecting amyloid pathology, regardless of cognitive status, as verified by amyloid PET across five clinical cohorts. These data support the potential of the pTau217 plasma immunoassay as a fully automated, high-throughput tool in study or clinical trial enrollment and routine clinical practice to aid in the timely diagnosis of AD and subsequent DMT treatment.

## AUTHOR CONTRIBUTIONS

Tobias Bittner, Derrek P. Hibar, Alina Bauer, and Christina Rabe wrote the manuscript. Derrek P. Hibar, Alina Bauer, and Christina Rabe performed the analysis. Niels Borlinghaus and Alexander Jethwa developed the Elecsys pTau217 immunoassay. Gwendlyn Kollmorgen, Henrik Zetterberg, and Kaj Blennow conducted the measurements with the Elecsys pTau217 immunoassay. Colin L. Masters and Reisa A. Sperling provided samples and data from the AIBL and A4 cohorts, respectively. All authors critically reviewed the analysis, data, and the manuscript.

## ACKNOWLEDGMENTS

We thank all the study participants and their families, the investigators, and site staff for their time and commitment. Henrik Zetterberg is a Wallenberg Scholar and a Distinguished Professor at the Swedish Research Council supported by grants from the Swedish Research Council (#2023-00356, #2022-01018 and #2019-02397), the European Union’s Horizon Europe research and innovation programme under grant agreement No 101053962, and Swedish State Support for Clinical Research (#ALFGBG-71320).

## DATA AVAILABILITY STATEMENT

Aggregated data are available on reasonable request from the corresponding author of this publication.

## ETHICAL STATEMENT

All studies included in this research were conducted in accordance with applicable ethical standards and regulatory guidelines. For A4 (NCT02008357), institutional review board approvals were obtained at each of the trial sites, and all participants provided written informed consent. The study was conducted in accordance with the International Council for Harmonisation (ICH) Good Clinical Practice (GCP) guidelines. The SKYLINE study (NCT05256134) was conducted in accordance with ICH-GCP guidelines and principles outlined in the Declaration of Helsinki. All study participants provided written, informed consent prior to any pre-screening, screening assessments, and/or intervention activities. The AIBL study and all subsequent protocol amends (SAGE Project ID Number: 2022/PID06188; SVHM Local Ref ID: HREC 028/06) received approval from institutional human research ethics committees of Austin Health, St Vincent’s Health, Hollywood Private Hospital, and Edith Cowan University. All volunteers provided written informed consent prior to participation. The study was conducted in accordance with the Helsinki Declaration of 1975. Both the CREAD (NCT02670083) and CREAD2 (NCT03114657) trials were conducted in accordance with ICH E6 GCP guidelines and the principles of the Declaration of Helsinki. All enrolled participants and their caregivers provided written informed consent prior to participation.

95% CI: 95% confidence interval
A4: Anti-Amyloid Treatment in Asymptomatic Alzheimer’s Disease
Aβ: amyloid-beta
AD: Alzheimer’s disease
AIBL: Australian Imaging Biomarkers and Lifestyle
*APOE* ε*4*: apolipoprotein E4
AUC: area under curve
BBBM: blood-based biomarker
BLOQ: number of samples/patients with pTau217 measurements below the limit of quantification
CDR-GS: Clinical Dementia Rating-Global Score
cgam: constrained generalized additive model
CI: cognitively impaired
CL: centiloid
CSF: cerebrospinal fluid
CU: cognitively unimpaired
DMT: disease-modifying therapy
DX: diagnosis
F: female
FDA: US Food and Drug Administration
LMCI: late mild cognitive impairment
M: male
MCI: mild cognitive impairment
Min: minimum
NEG: negative
NPV: negative predictive value
PET: positron emission tomography
POS: positive
PPV: positive predictive value
pTau217: tau phosphorylated at threonine 217
ROC: receiver operating characteristic
SD: standard deviation
SF: screen fails
SUVr: standardized uptake value ratio
USA: United States of America

