## Supplementary material for "Performance of Elecsys pTau217 plasma immunoassay to detect brain amyloid pathology": Suppl Table 1

### SUPPLEMENTARY MATERIALS

**Supplementary Table 1:** Summary of baseline characteristics of the overall population in participants with available amyloid PET visual read measurements

|  |  | **A4 CU (*N* = 4486)** | **SKYLINE CU  (*N* = 352)** | **AIBL** | | **CREAD CI (*N* = 783)** | **CREAD2 CI (*N* = 774)** | **Overall** | |
| --- | --- | --- | --- | --- | --- | --- | --- | --- | --- |
|  |  |  |  | **CI (*N* = 163)** | **CU (*N* = 694)** |  |  | **CI (*N* = 1720)** | **CU (*N* = 5532)** |
| Age, years | Mean (SD) | 70.8 (4.66) | 70.2 (5.31) | 72.5 (7.04) | 71.3 (5.08) | 71.6 (7.91) | 71.4 (7.53) | 71.6 (7.66) | 70.9 (4.77) |
|  | Median  (Min, Max) | 70.0  (64.0, 85.0) | 70.0  (60.0, 81.0) | 73.0  (51.0, 86.0) | 70.0  (59.0, 90.0) | 73.0  (50.0, 86.0) | 72.0  (50.0, 85.0) | 73.0  (50.0, 86.0) | 70.0  (59.0, 90.0) |
| Sex, *n* (%) | F | 2663 (59.4) | 229 (65.1) | 80 (49.1) | 395 (56.9) | 455 (58.1) | 431 (55.7) | 966 (56.2) | 3287 (59.4) |
|  | M | 1823 (40.6) | 123 (34.9) | 83 (50.9) | 299 (43.1) | 328 (41.9) | 343 (44.3) | 754 (43.8) | 2245 (40.6) |
| Race, *n* (%) | American Indian or Alaska Native | 9 (0.2) | 0 | 0 | 0 | 0 | 1 (0.1) | 1 (0.1) | 9 (0.2) |
|  | Asian | 170 (3.8) | 7 (2.0) | 0 | 0 | 63 (8.0) | 102 (13.2) | 165 (9.6) | 177 (3.2) |
|  | Black or African American | 159 (3.5) | 27 (7.7) | 0 | 0 | 19 (2.4) | 10 (1.3) | 29 (1.7) | 186 (3.4) |
|  | Multiple | 27 (0.6) | 0 | 0 | 0 | 1 (0.1) | 2 (0.3) | 3 (0.2) | 27 (0.5) |
|  | Native Hawaiian or Other Pacific Islander | 2 (0.0) | 0 | 0 | 0 | 0 | 0 | 0 | 2 (0.0) |
|  | Unknown | 26 (0.6) | 64 (18.2) | 0 | 0 | 15 (1.9) | 11 (1.4) | 26 (1.5) | 90 (1.6) |
|  | White | 4093 (91.2) | 247 (70.2) | 163 (100) | 694 (100) | 685 (87.5) | 648 (83.7) | 1496 (87.0) | 5034 (91.0) |
|  | Other | 0 | 7 (2.0) | 0 | 0 | 0 | 0 | 0 | 7 (0.1) |
| Ethnicity,  *n* (%) | Hispanic or Latino | 142 (3.2) | 62 (17.6) | 0 | 0 | 69 (8.8) | 38 (4.9) | 107 (6.2) | 204 (3.7) |
|  | Not Hispanic or Latino | 4309 (96.1) | 16 (4.5) | 0 | 0 | 702 (89.7) | 718 (92.8) | 1420 (82.6) | 4325 (78.2) |
|  | Unknown | 35 (0.8) | 266 (75.6) | 0 | 0 | 6 (0.8) | 6 (0.8) | 12 (0.7) | 301 (5.4) |
|  | Not reported | 0 | 8 (2.3) | 0 | 0 | 6 (0.8) | 12 (1.6) | 18 (1.0) | 8 (0.1) |
|  | Missing | 0 | 0 | 163 (100) | 694 (100) | 0 | 0 | 163 (9.5) | 694 (12.5) |
| *APOE ε4*,  *n* (%) | 0 ε4 | 2894 (64.5) | 228 (64.8) | 68 (41.7) | 489 (70.5) | 290 (37.0) | 310 (40.1) | 668 (38.8) | 3611 (65.3) |
|  | 1 ε4 | 1415 (31.5) | 108 (30.7) | 67 (41.1) | 175 (25.2) | 383 (48.9) | 340 (43.9) | 790 (45.9) | 1698 (30.7) |
|  | 2 ε4 | 141 (3.1) | 13 (3.7) | 23 (14.1) | 23 (3.3) | 108 (13.8) | 124 (16.0) | 255 (14.8) | 177 (3.2) |
|  | Missing | 36 (0.8) | 3 (0.9) | 5 (3.1) | 7 (1.0) | 2 (0.3) | 0 | 7 (0.4) | 46 (0.8) |
| Diagnosis,  *n* (%) | AD | 0 | 0 | 60 (36.8) | 0 | 0 | 0 | 60 (3.5) | 0 |
|  | SF/LMCI/AD | 0 | 0 | 0 | 0 | 783 (100) | 774 (100) | 1557 (90.5) | 0 |
|  | MCI | 0 | 0 | 103 (63.2) | 0 | 0 | 0 | 103 (6.0) | 0 |
|  | CN | 4486 (100) | 352 (100) | 0 | 694 (100) | 0 | 0 | 0 | 5532 (100) |
| CDR-GS,  *n* (%) | 0 | 4484 (100) | 343 (97.4) | 9 (5.5) | 665 (95.8) | 0 | 0 | 9 (0.5) | 5492 (99.3) |
|  | 0.5 | 2 (0.0) | 0 | 119 (73.0) | 28 (4.0) | 567 (72.4) | 587 (75.8) | 1273 (74.0) | 30 (0.5) |
|  | 1 | 0 | 0 | 28 (17.2) | 0 | 216 (27.6) | 186 (24.0) | 430 (25.0) | 0 |
|  | 2 | 0 | 0 | 6 (3.7) | 0 | 0 | 0 | 6 (0.3) | 0 |
|  | 3 | 0 | 0 | 1 (0.6) | 0 | 0 | 0 | 1 (0.1) | 0 |
|  | Missing | 0 | 9 (2.6) | 0 | 1 (0.1) | 0 | 1 (0.1) | 1 (0.1) | 10 (0.2) |
| Centiloid ≥ 24, *n* (%) | NEG | 3040 (67.8) | 244 (69.3) | 42 (25.8) | 526 (75.8) | 172 (22.0) | 152 (19.6) | 366 (21.3) | 3810 (68.9) |
|  | POS | 1446 (32.2) | 98 (27.8) | 121 (74.2) | 168 (24.2) | 565 (72.2) | 538 (69.5) | 1224 (71.2) | 1712 (30.9) |
|  | Missing | 0 | 10 (2.8) | 0 | 0 | 46 (5.9) | 84 (10.9) | 130 (7.6) | 10 (0.2) |
| Centiloid ≥ 40, *n* (%) | NEG | 3451 (76.9) | 263 (74.7) | 52 (31.9) | 575 (82.9) | 225 (28.7) | 216 (27.9) | 493 (28.7) | 4289 (77.5) |
|  | POS | 1035 (23.1) | 79 (22.4) | 111 (68.1) | 119 (17.1) | 512 (65.4) | 474 (61.2) | 1097 (63.8) | 1233 (22.3) |
|  | Missing | 0 | 10 (2.8) | 0 | 0 | 46 (5.9) | 84 (10.9) | 130 (7.6) | 10 (0.2) |
| Visual read, *n* (%) | NEG | 3813 (85.0) | 311 (88.4) | 58 (35.6) | 610 (87.9) | 233 (29.8) | 226 (29.2) | 517 (30.1) | 4734 (85.6) |
|  | POS | 673 (15.0) | 41 (11.6) | 105 (64.4) | 84 (12.1) | 550 (70.2) | 548 (70.8) | 1203 (69.9) | 798 (14.4) |

Abbreviations: A4, Anti-Amyloid Treatment in Asymptomatic Alzheimer’s Disease; AD, Alzheimer’s disease;
AIBL, Australian Imaging Biomarkers and Lifestyle; *APOE ε4*, apolipoprotein E4; CDR-GS, Clinical Dementia Rating-Global Score; CI, cognitively impaired; CN, cognitively normal; CU, cognitively unimpaired; F, female;
LMCI, late mild cognitive impairment; MCI, mild cognitive impairment; M, male; Max, maximum; Min, minimum,
NEG, negative, PET, positron emission tomography; POS, positive; SD, standard deviation; SF, screen fails.
