## Supplementary material for "Performance of Elecsys pTau217 plasma immunoassay to detect brain amyloid pathology": Suppl Figures 1&2

**Supplementary Figure 1:** ROC analyses for plasma pTau217 measurements with respect to amyloid PET visual read status in (A) cognitively impaired and (B) cognitively unimpaired individuals.

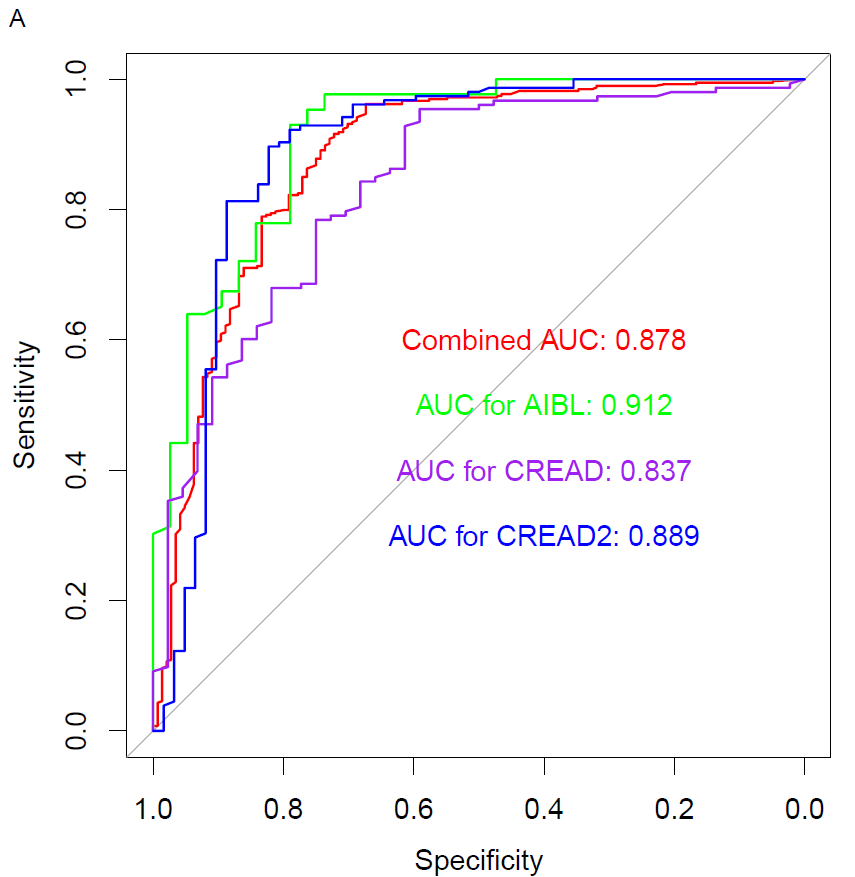

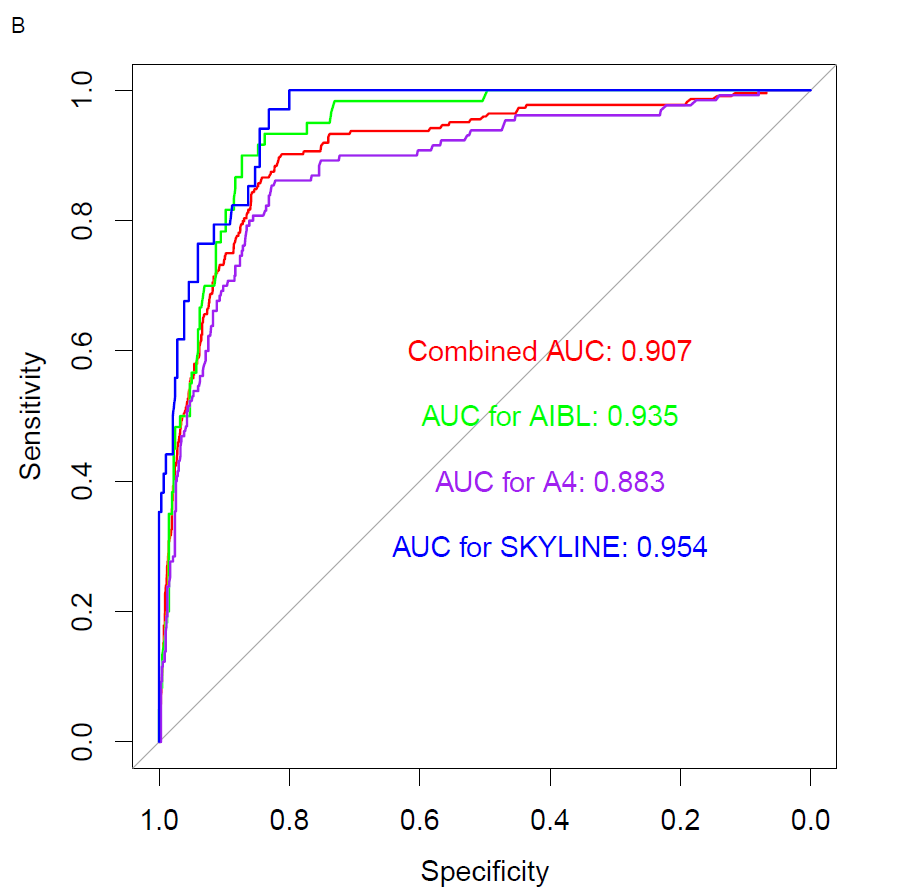

Abbreviations: A4, Anti-Amyloid Treatment in Asymptomatic Alzheimer’s Disease;
AIBL, Australian Imaging Biomarkers and Lifestyle; AUC, area under curve;
PET, positron emission tomography; pTau217, tau phosphorylated at threonine 217; ROC, receiver operating characteristic.

**Supplementary Figure 2:** Centiloid classifications vs amyloid PET visual read status
by diagnosis across the five clinical cohorts.

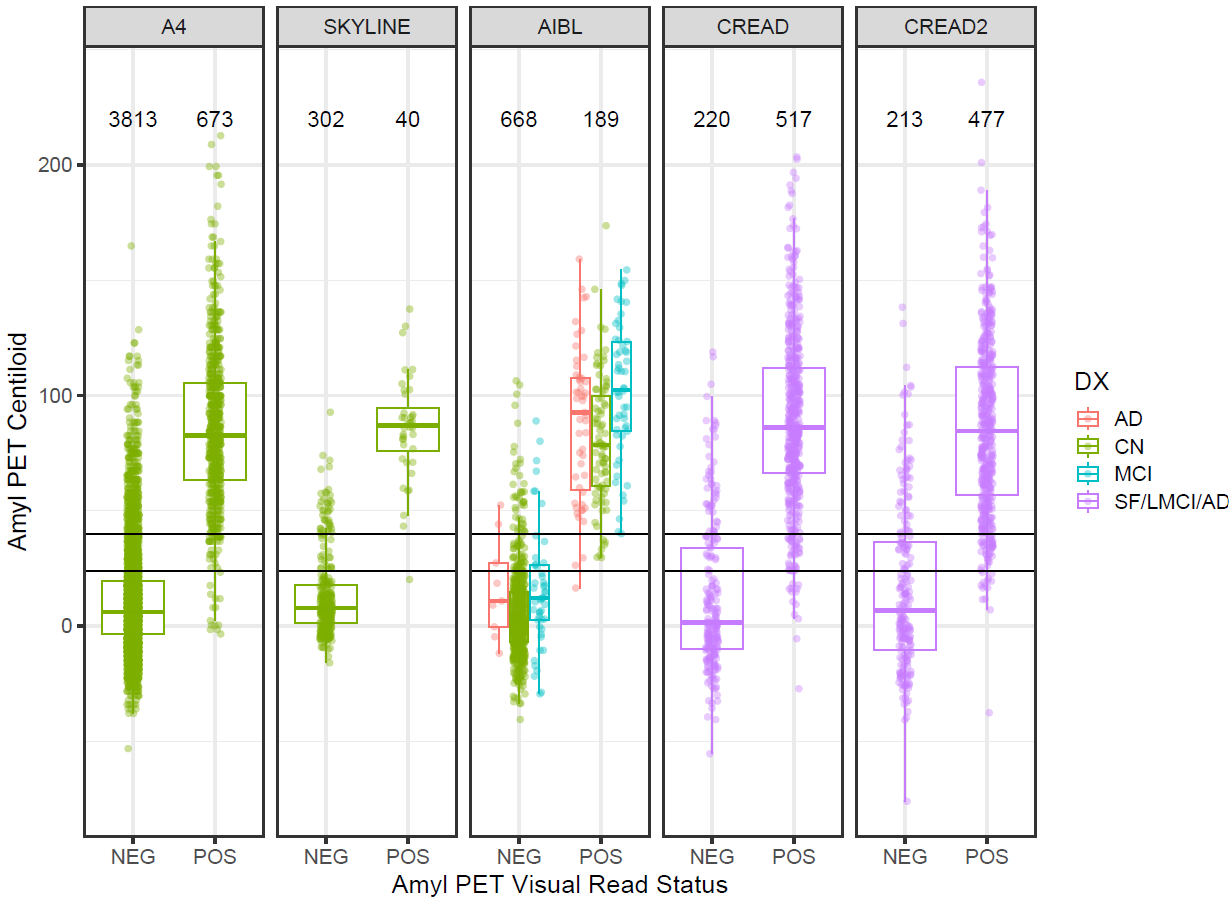

Horizontal black lines indicate centiloid cutoffs of ≥ 24 and ≥ 40.

Abbreviations: A4, Anti-Amyloid Treatment in Asymptomatic Alzheimer’s Disease;
AD, Alzheimer’s disease; AIBL, Australian Imaging Biomarkers and Lifestyle;
Amyl, amyloid; CN, cognitively normal; DX, diagnosis; LMCI, late mild cognitive impairment; MCI, mild cognitive impairment; NEG, negative; POS, positive;
PET, positron emission tomography; SF, screen fails.
